# Epidemiological, Clinical, and Diagnostic Characteristics of a Large-Scale Upsurge of Dengue in the Rohingya Refugee Camps and Host Communities in Cox’s Bazar, Bangladesh, 2021 to 2024: A Retrospective Study

**DOI:** 10.64898/2026.03.27.26349486

**Authors:** Charls Erik Halder, Md Abeed Hasan, Emmanuel Roba Soma, James Charles Okello, Md Mostafizur Rahman, Partho Pratim Das, U Maung Prue, Dickson Wafula Barasa, Md Atiquzzaman, Md Sobuz Hosen, Saddam Hossain Shagar, Elizabeth Yie Chuen Chong, Debashish Paul, S M Niaz Mowla, Mohammadul Hoque, Abu Toha MHR Bhuiyan, Md Farhad Hussain

## Abstract

**Background:** Dengue emerged as a new public health threat in the Rohingya refugee camps in Cox’s Bazar, Bangladesh, in 2021 and expanded into large-scale upsurges in subsequent years. Evidence on dengue epidemiology and clinical presentation in protracted refugee settings remains limited, despite the need for stronger outbreak preparedness and case management in these contexts.

**Objectives:** To describe the epidemiological, clinical, and diagnostic characteristics of the dengue upsurge among Rohingya refugees and surrounding host communities in Cox’s Bazar, Bangladesh, and to identify predictors of inpatient admission and diagnostic positivity patterns.

**Methods:** This retrospective observational study used anonymized surveillance data from the International Organization for Migration (IOM) dengue patient database. Rapid diagnostic test (RDT)–confirmed dengue cases identified across 36 IOM-supported health facilities in Ukhiya and Teknaf between 1 October 2021 and 31 December 2024 were included. Demographic, epidemiological, clinical, and laboratory variables were summarized using descriptive statistics. Weekly incidence was aggregated by epidemiological week and calendar year, and epidemic growth and decay phases were modelled using phase-specific Poisson regression. Multivariable logistic regression was used to identify predictors of inpatient admission and to examine associations between delay in presentation and RDT positivity patterns, adjusting for age and sex.

**Results:** A total of 35,581 RDT-confirmed dengue cases were reported, of which 90.2% occurred among Rohingya refugees. The median age was 17 years (IQR 7–30), and 46.0% of cases were among children aged 0–14 years. Annual caseload increased from 1,011 in 2021 to 11,752 in 2022, 10,669 in 2023 and 12,149 in 2024, with seasonal peaks during the monsoon period and progressively later peaks and longer epidemic tails over time. Poisson models showed decreasing growth rates across years (r=0.449 in 2021 to r=0.091 in 2024) with increasing doubling times, while decay rates remained broadly comparable (halving time ∼4.4–6.0 weeks). Overall, 8.0% of cases required inpatient admission, 1.3% were referred, and four deaths were reported (case fatality <0.1%).

In multivariable analysis, inpatient admission was associated with older age (≥60 vs 0–14: aOR 2.31), delayed presentation (aOR 1.06 per day), refugee status (aOR 1.39), presence of any World Health Organization (WHO) warning sign (aOR 26.60), low systolic BP (aOR 2.84) and chronic co-morbidity (aOR 6.07). In addition, males had lower odds of admission than females (aOR 0.88). NS1 antigen alone was positive in 62.1% of cases, IgM alone in 33.6%, and dual positivity in 4.3%. Longer delay from symptom onset to presentation was strongly associated with IgM-only positivity compared with NS1-only positivity (adjusted models controlling for age and sex).

**Conclusion:** Sustained dengue preparedness is required in Cox’s Bazar, including strengthened surveillance, community-based early referral, targeted monitoring of high-risk groups, environmental vector control, and phase-appropriate use of NS1 and IgM/IgG diagnostics to reduce missed diagnoses and prevent progression to severe disease. These findings highlight the need for a policy shift from episodic outbreak response toward sustained dengue preparedness in humanitarian settings, including strengthened surveillance systems, integrated diagnostic strategies, community-based early referral, and coordinated vector control interventions.

## INTRODUCTION

Rohingya refugees have experienced multiple instances of forced displacement into Bangladesh as a result of conflict and persecution since 1978 with its’ largest displacement of the Rohingyas from Myanmar to Bangladesh occurring in August 2017 (1,2). As of 31st December 2025, approximately 1,148,571 Rohingya refugees were residing in 33 overcrowded camps in Ukhiya and Teknaf Upazilas of Cox’s Bazar (3,4). The Rohingya refugees in Cox’s Bazar continue to face challenging living conditions characterized by overcrowded camps, limited access to essential resources and services, and increased vulnerability to natural and man-made disasters, like floods and landslides related to heavy monsoons rainfall, fire incidents and cyclones (1). The crowded living arrangements with fragile shelters, inadequate water, sanitation, and hygiene (WASH) facilities and practices, coupled with intense monsoons rainfall in both the refugee camps and neighboring host communities, elevate their vulnerability to infectious diseases, frequently leading to disease outbreaks (1). Since 2017, these areas have witnessed various episodes of infectious disease outbreaks or upsurges, including diphtheria, measles, COVID-19, and Acute Watery Diarrhea (AWD)/cholera (1,6). Dengue appeared as a new threat for the Rohingya refugees from October 2021 which expanded as massive outbreaks in 2022 and 2023 (7–9).

Dengue is a viral infection caused by the Dengue virus (DENV), transmitted to humans through the bite of infected mosquitoes (10). Despite DENV having four serotypes (DENV-1, DENV-2, DENV-3, DENV-4), infection with one serotype provides long-term immunity to only that serotype but not the others, with sequential infections raising the risk of severe dengue (11). A patient with dengue can manifest with a spectrum of symptoms, ranging from mild to severe ones (10,11). Common symptoms reported for dengue include high fever (40°C/104°F), severe headache, retro-orbital pain (pain behind the eyes), muscle and joint pains, nausea, vomiting, swollen glands, and rash (10,12). Severe dengue symptoms often manifest after the fever has subsided and may include severe abdominal pain, persistent vomiting, rapid breathing, bleeding gums or nose, fatigue, restlessness, blood in vomit or stool, extreme thirst, pale and cold skin, and feeling weak (10,11). Epidemiological and clinical characteristics of dengue have been described in several studies conducted over the past two decades, particularly from India, Myanmar, Thailand, and Malaysia (13–16). However, most of the studies described the characteristics of dengue in urban setting or whole country setting (13,17). Even the studies in Bangladesh also described the characteristics either mainly from the urban setting or based on the data from the whole country (17–19).

A few studies have been conducted on zoonotic diseases and applied research among displacement populations such as in internally displaced populations (IDP) camps in Sri Lanka and other global settings, yet a significant literature gap on the epidemiological and clinical characteristics of dengue still exists, especially within the Rohingya humanitarian context. This understanding is critical for projecting the nature of upcoming outbreaks and ensuring proper prevention, preparedness, and response strategies (1,8). Previous studies in the Rohingya camps have also highlighted important gaps in preventive practices, community engagement, and risk communication during infectious disease responses, which remain highly relevant for dengue preparedness and response in this setting (5,6). Moreover, there has been concerns in recent years that dengue is being presented atypically with new signs, symptoms and diagnostic characteristics, for instance, presentation with cough, diarrhea and gastrointestinal manifestation (20–22).

The Rohingya refugee camps in Bangladesh constitute a distinctive environment, hosting more than one million refugees living in improvised and overcrowded conditions marked by limited waste disposal, poor drainage systems, and makeshift shelters constructed from bamboo and tarpaulins (1). An investigation into epidemiological factors is imperative for anticipating the likelihood, magnitude, duration, and peak timing of dengue outbreaks, facilitating the timely implementation of preventive measures and the establishment of preparedness and response plans (11,23). Furthermore, an examination of the epidemiological and clinical features of dengue within this displaced setting is essential for formulating outbreak preparedness and response plans, strengthening case management measures while ensuring prompt implementation of risk communication and community engagement initiatives, and the forecasting and strategic prepositioning of medical commodities and supplies (11,24).

The objective of this study was to describe the epidemiological, clinical and diagnostic characteristics of large-scale dengue upsurge in the Rohingya refugee camps as well as in the surrounding host communities in Cox’s Bazar, Bangladesh. To be more specific, this study was designed to characterize transmission dynamics, clinical presentations, severity of the disease, pattern of presentation in health facilities, and diagnostic profiles of dengue cases in this protracted humanitarian setting. This study aimed to contribute to the evidence relevant to dengue outbreak preparedness, surveillance, and response strategies in displaced populations’ settings.

## METHODOLOGY

### STUDY DESIGN

This was a retrospective observational study that utilized de-identified and anonymized data from the International Organization for Migration’s (IOM) dengue patient database in Cox’s Bazar, Bangladesh.

### STUDY SETTING AND POPULATION

The study was implemented in Rohingya refugee camps in Ukhiya and Teknaf in Cox’s Bazar. The study population included all individuals who were diagnosed with dengue and received care across 36 IOM-supported health facilities in Ukhiya and Teknaf, Bangladesh, between October 1, 2021, and December 31, 2024. Dengue was first detected in this displacement setting in 2021 and subsequently evolved into multiple large-scale outbreaks, prompting a substantial response from IOM across the Rohingya camps. Therefore, these camps were selected for this study to gather an in-depth understanding of the outbreak.

### DATA SOURCE

#### Dengue data collection and case definition

De-identified and anonymized demographic, epidemiological, and clinical data pertaining to confirmed dengue cases irrespective of age among Rohingya refugees were retrieved from the International Organization for Migration’s (IOM) dengue patient database spanning the period from 1^st^ October 2021 to 31^st^ December 2024. All dengue cases were screened and identified across 36 IOM supported facilities in Ukhiya and Teknaf. The following case definitions were used for screening and diagnosis of the patients as per the national guideline for management of dengue syndrome in Bangladesh (24).

- Suspected dengue: Acute febrile illness with or without non-specific signs and symptoms.
- Confirmed dengue: Case patients reported as positive for NS1 antigen or IgM by rapid diagnostic tests

The health facilities collected daily data for demographic (e.g. age, sex, camp location), epidemiological clinical (e.g. symptoms, severity, complications, hospitalization, hospital stay, and referral), and laboratory findings (e.g. RDT result) of the cases using standard Case Report Form and Inpatient Admission Notification Form (in case of inpatient admission). All data were reported electronically using Kobo Toolbox and synchronized at the central database. Data was verified and cleaned centrally on a daily basis, then exported to an excel spreadsheet for further analysis.

#### Clinical Management

Out of 36 IOM-supported health facilities, 22 were health posts that operated only during daytime (8.00 AM to 4.00 PM), while 12 were 24/7 primary healthcare centers (PHCCs) with inpatient capacity, and 2 were integrated infectious disease treatment centers (IDTC) dedicated for management and admission of moderate to severe cases. The management of dengue cases followed the Bangladesh national treatment guidelines, which classified cases into groups A, B, and C corresponding to mild, moderate, and severe categories (24). This classification aligns with the WHO’s revised classification (2009) for dengue hemorrhagic fever (DHF), which categorized dengue into three groups (A-B-C) to streamline case management based on disease severity (Figure 1). Group A patients were managed at all three levels of health facilities. IDTCs admitted Group B patients as well as patients with social factors (e.g. living far from the facility, living alone). IDTCs admitted patients referred from health posts or PHCCs (if the bed capacity is saturated). Group C patients were referred to District Sadar Hospital for initial stabilization. A dispatch and referral unit monitored real-time bed capacity across the health facilities, coordinated referrals, and dispatched ambulances. In addition, a network of Community Health Workers (CHWs) conducted daily follow-up of dengue cases managed in the outpatient departments (OPD) of the health facilities and monitored patients for warning signs using a standardized dengue community follow-up checklist.

**Figure 1.**
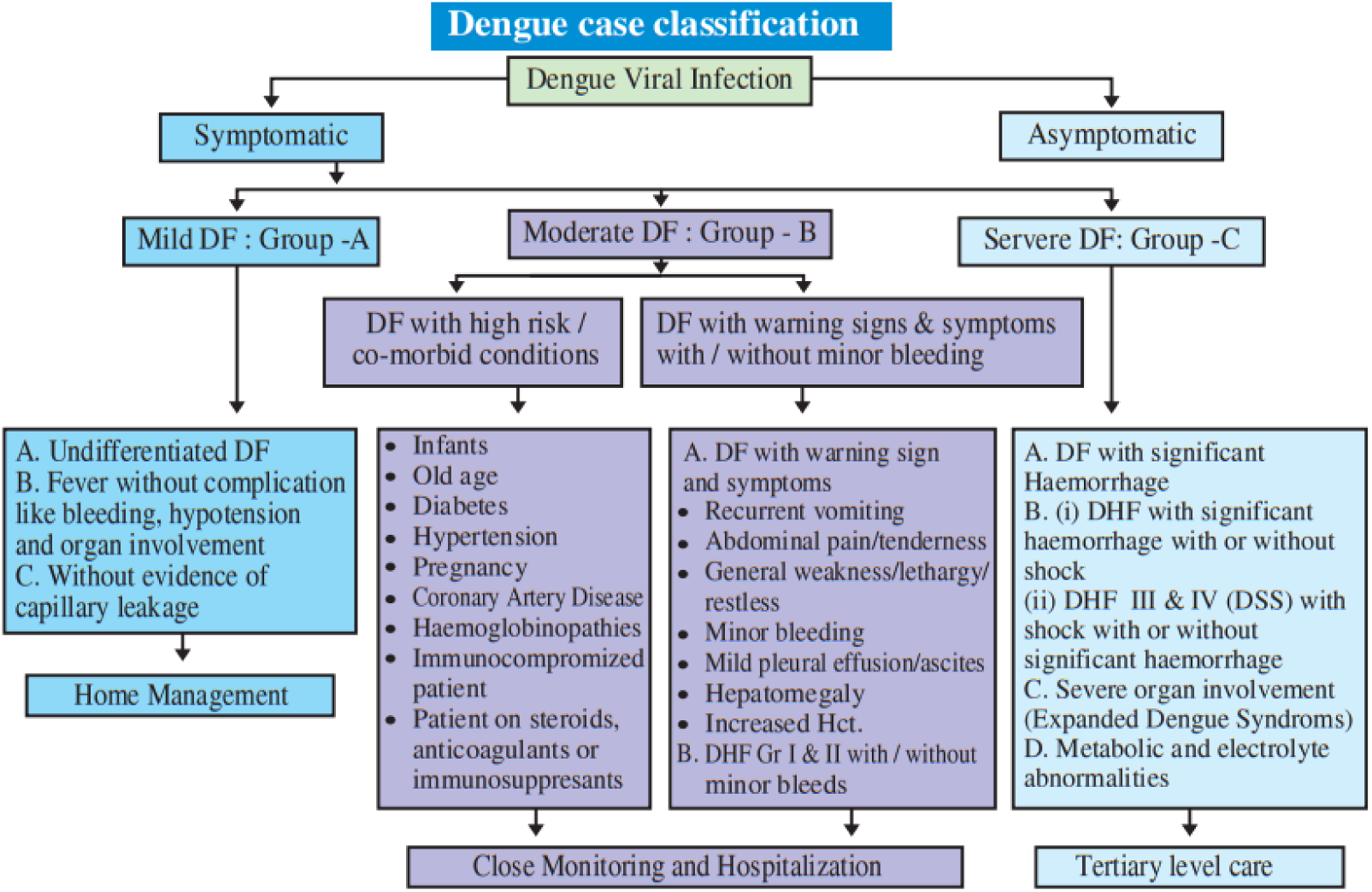
Dengue case classification used in clinical management (Groups A–C) (24).

#### Diagnostics

NS1 antigen and IgM/IgG RDT kits were used across all facilities to diagnose dengue. Venous blood samples were collected from the patients for the tests. The NS1 antigen RDT was administered for patients in the early or febrile phase, while the IgM/IgG antibody RDT was utilized if the patient presented in the late or convalescence phase. This approach aligned with findings from systematic reviews, demonstrating the high sensitivity of combining NS1 and IgM tests during the respective phases of dengue. In instances where differentiation between febrile and convalescent phases was challenging, or diagnostic uncertainty persisted with a single RDT result, both tests were conducted.

## Data Analysis

Demographic, epidemiological, and clinical characteristics were analyzed using descriptive statistics. These included frequencies, percentages, means, and medians, as appropriate. Differences in proportions across demographic categories were assessed using chi-square tests. Attack rate ratios and corresponding p-values were estimated using Poisson regression, with the relevant reference category specified for each comparison. Demographic variables were age, sex, population type (refugee or host community) as well as the geographical location. Clinical characteristics included symptoms during presentation, vital signs, presence of dengue warning signs as per WHO, presence of co-morbidities including pregnancy status and non-communicable diseases. This also captured the delay from onset of symptoms to presentation at a health facility. Diagnostic variables were RDT results (NS1 antigen and IgM). The standardized dengue case report form used for routine data collection is provided in S2 File.

Weekly dengue incidence was aggregated by epidemiological week and calendar year based on the date of clinical examination. Epidemic curves were constructed for each year from 2021 to 2024. The peak week for each annual epidemic was determined by identifying the epidemiological week with the maximum observed weekly case count. Each epidemic was then divided into two distinct phases, a growth phase; defined as weeks from epidemic onset up to and including the peak week, and a decay phase; defined as weeks following the peak. Phase-specific exponential trends were modelled using Poisson regression with a log link function, where weekly case counts were regressed against epidemiological week within each phase. The estimated regression coefficient for epidemiological week (r) represents the weekly exponential growth (r > 0) or decay (r < 0) rate. Ninety-five percent confidence intervals were obtained from model-based standard errors. Doubling time during growth phases was calculated as ln(2)/r, while halving time during decay phases was calculated as ln(2)/|r|. Phase-specific fitted values were overlaid on observed weekly incidence to visually assess model fit. Analyses were restricted to completed epidemic years (2021–2024) to avoid bias from incomplete epidemic trajectories.

Delay in presentation was analysed as a continuous variable and also compared across RDT positivity patterns using non-parametric methods. The distribution of RDT positivity patterns by delay from symptom onset to presentation is illustrated in S1 Figure. Differences in delay across RDT categories were assessed using the Kruskal–Wallis test. Multivariable logistic regression models were used to examine the association between delay in presentation and RDT positivity patterns. And the reference category was NS1 antigen–only positivity. These models were adjusted for age and sex. Results were reported as adjusted odds ratios (aORs) with 95% confidence intervals.

To identify the predictors of inpatient admission, univariable and multivariable logistic regression analyses were performed. Covariates were selected a priori based on clinical relevance and WHO dengue severity criteria. This included age group, sex, delay in presentation, population type, fever at presentation, systolic and diastolic blood pressure (BP), presence of any warning sign as per WHO, pregnancy, chronic co-morbidities, and previous dengue infection. Adjusted odds ratios with 95% confidence intervals were estimated using maximum likelihood methods.

Model performance was evaluated using likelihood ratio tests comparing the final models with null models. Discrimination was assessed using the area under the receiver operating characteristic curve (AUC). And calibration was examined using decile-based calibration plots and the Hosmer–Lemeshow goodness-of-fit test. Multicollinearity among predictors was assessed using variance inflation factors. All analyses were conducted using R statistical software (version 4.4.2; R Core Team, 2024).

### Research Governance

This research aligned with the global strategy and priorities of IOM in relation to ensuring access to healthcare to save lives and protect the displaced populations. All existing literature, scientific evidence and ethical considerations were taken into account while designing the research protocol. Existing public health experts of IOM in the Migration Health Division, Cox’s Bazar took part in the research. While the first researcher (as mentioned under the research team) led the technical aspects of the research including protocol development, ethical consideration, training, data collection and analysis, and reporting, the second researcher led the research coordination, monitoring, budget allocation, and risk management. Other researchers took part in data collection, cleaning, analysis and review. All research team members (CV attached) possessed the relevant professional qualifications to contribute to the research study. Both first and second researchers did regular monitoring of timely implementation and quality of every step of the research. Training were organized for research team members on ethics, data collection, data protection, analysis and presentation.

### Limitations

The study did not include genotyping of the circulating dengue virus strains. Evidence from recent studies suggested that the emergence of the DENV-3 serotype, which was undetected for a long time, had been associated with increased clinical severity in dengue outbreaks (25). The absence of genotyping in this study may limited our understanding of how changes in dengue serotypes contribute to the observed clinical manifestations and the severity of outbreaks in this population. In addition, although surveillance data were available for the full year 2021, dengue transmission was first detected only in October 2021. Comparisons of epidemic timing and trajectory between 2021 and the full calendar years of 2022 to 2024 should therefore be interpreted cautiously.

### Ethical Consideration

This study was conducted under a pre-approved protocol for secondary analysis of outbreak-related health and programme data. Ethical approval was obtained from the Ethical Review Board of Cox’s Bazar Medical College Hospital (CoxMC/2023/016). Administrative permission to use outbreak-response data was granted by the Office of the Civil Surgeon and the Refugee Relief and Repatriation Commissioner (RRRC). Internal approval was also obtained from the International Organization for Migration (IOM) Migration Health Research Unit for the study and retrospective use of programme data.

Epidemiological and clinical case data were extracted from routine outbreak surveillance and clinical reporting databases used during the dengue response. These data were collected as part of public health outbreak response and health service delivery activities between 1 October 2021 and 31 December 2024 and subsequently used for research purposes under the approved secondary-use protocol. Anonymized surveillance database was accessed for research purposes between 01 August 2025 and 15 September 2025. All individual-level data were de-identified and anonymized prior to analysis, and only aggregate findings are reported. At no point did the investigators have access to direct or indirect patient identifiers. Given the retrospective nature of the study and the exclusive use of anonymized data, individual informed consent was not required.

## RESULT

### Demographics

A total of 35,581 rapid diagnostic test confirmed dengue positive cases were reported from the selected health facilities in the Rohingya refugee camps, of which 90.2% of the cases were among Rohingya refugees and rest were among local Bangladeshi communities. The annual attack rate ranged from 14.6 to 175.6 per 10,000 person-years between 2021 and 2024, with a cumulative attack rate of 514.3 cases per 10,000 person-years over the study period (Table 1).

**Table 1.**
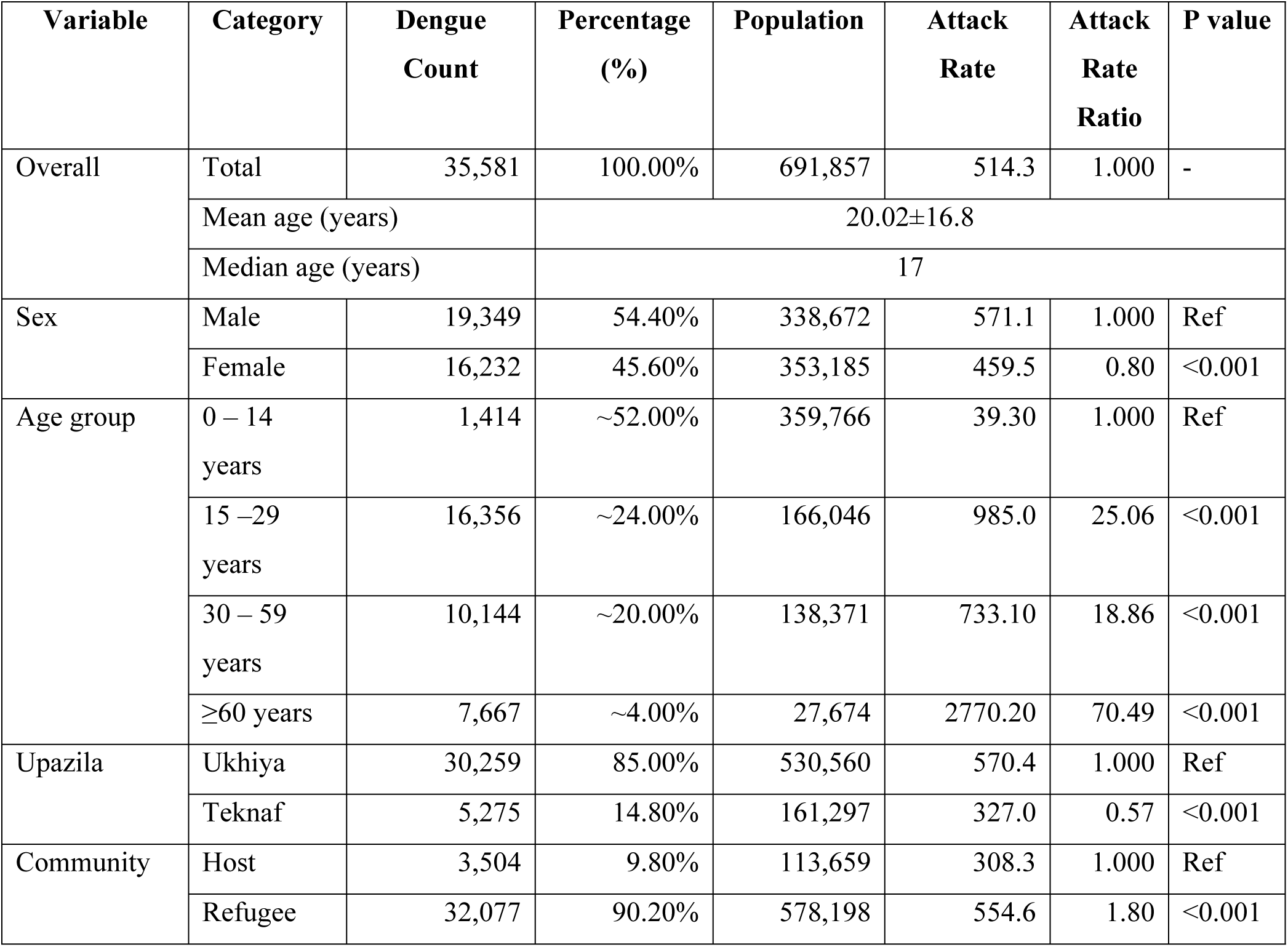
Distribution of dengue cases and attack rates by demographic characteristics across 2021 to 2024 (N = 35,581)

The median age of the cases were 17 years (IQR 7 – 30 years). Nearly half of all reported dengue cases occurred among children aged 0–14 years. The majority of cases were documented in Ukhiya Upazila (85.0%), followed by Teknaf (14.8%). A higher proportion of cases was observed among males compared with females (54.4% vs. 45.6%), indicating a modest male predominance in dengue incidence (Table 2).

**Table 2.** Demographic characteristics of dengue cases at presentation across 2021 to 2024 (N = 35,581)

| Characteristic | Category | n (%) or value |
| --- | --- | --- |
| **Overall caseload** | Total cases | 35,581 (100.0) |
| **Sex** | Female | 16,232 (45.6) |
|  | Male | 19,349 (54.4) |
| **Age (years)** | Mean (SD) | 20.2 (16.8) |
|  | Median (IQR) | 17 (7–30) |
|  | Range | 0.003–117 |
| **Age group distribution** | 0–14 years | 16,371 (46.0) |
|  | 15–29 years | 10,141 (28.5) |
|  | 30–59 years | 7,655 (21.5) |
|  | ≥60 years | 1,414 (4.0) |
| **Population type** | Host community | 3,504 (9.8) |
|  | Refugee | 32,077 (90.2) |
| **Geographic distribution (Upazila)** | Ukhiya | 30,259 (85.0) |
|  | Teknaf | 5,275 (14.8) |
|  | Ramu | 25 (0.1) |
|  | Cox’s Bazar Sadar | 22 (0.1) |

### Epidemiological characteristics

Figure 2 illustrates the weekly trends in dengue cases by year, highlighting variations in epidemic timing, with progressively later peaks and extended epidemic tails over successive years. The initial outbreak was reported in October 2021, corresponding to the lowest overall case count (1,011), which increased in subsequent years (11,752 in 2022; 10,669 in 2023; and 12,149 in 2024). In all years, case numbers remained relatively low during the early epidemiological weeks, followed by a marked seasonal increase during the Monsoon period, with peaks occurring between weeks 25 and 38, varying by year. Notably, 2022 saw an early surge with a pronounced peak around weeks 25–26, while 2023 displayed a delayed and less acute peak near week 30. In contrast, 2024 experienced a prolonged epidemic characterized by a later peak during weeks 34–36 and sustained elevated incidence prior to decline.

**Figure 2.**
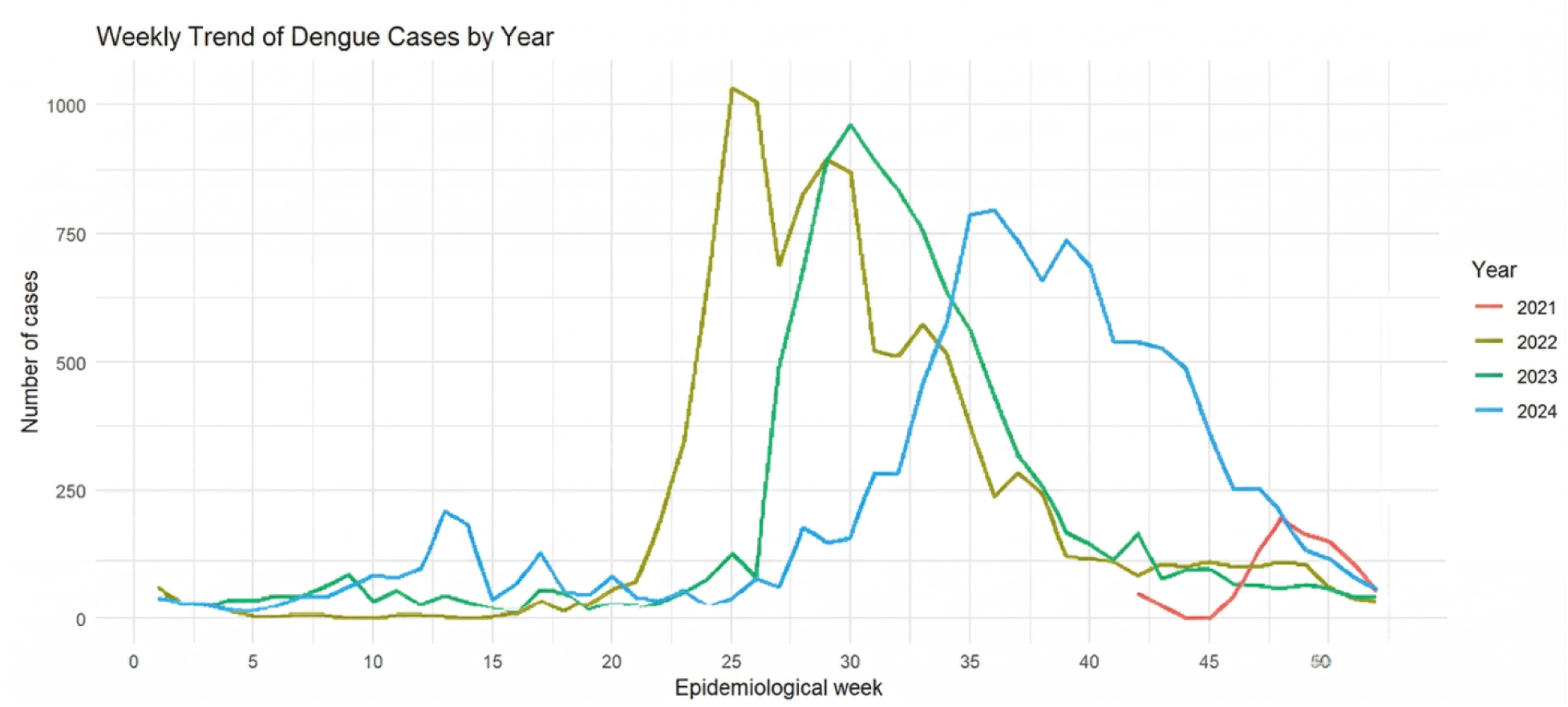
Weekly dengue cases by epidemiological week and year (2021–2024).

A phase-specific Poisson regression analysis demonstrates clear growth and decay phases in dengue transmission across all years (2021 to 2024) with notable variations in epidemic expansion and decline (Table 3, Figure 3). In October 2021, transmission began with a rapid surge and sharp decline, characterized by the fastest growth rate (r = 0.449; 95% CI: 0.394–0.506) and the shortest doubling time of 1.54 weeks. This was followed by a significant decline (r = −0.285; 95% CI: −0.364 to −0.208), with a halving time of 2.43 weeks. As surveillance began in October 2021, the 2021 growth and decay estimates should be interpreted cautiously, as the shorter observation window may have influenced the observed epidemic trajectory. From 2022 to 2024, epidemic growth became progressively slower, with reduced growth rates (r = 0.299 in 2022; 0.149 in 2023; 0.091 in 2024) and corresponding increase in doubling times, lengthened from 2.32 to 7.61 weeks. In contrast, post-peak decay rates remained generally comparable across years (ranging from −0.116 to −0.159), with halving times between 4.37 and 5.97 weeks, indicating more prolonged epidemic persistence despite attenuated transmission intensity.

**Figure 3.**
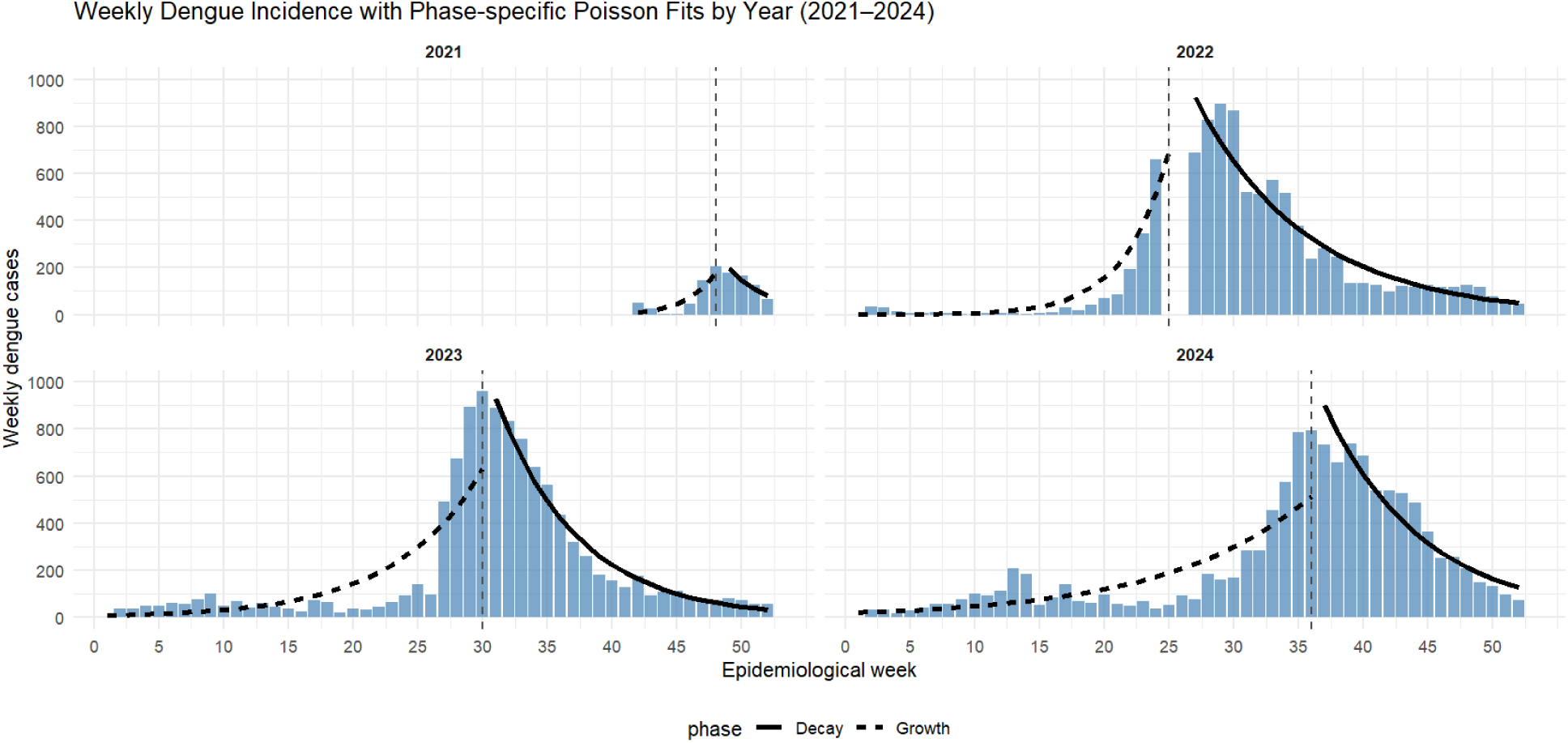
Weekly dengue incidence with phase-specific Poisson regression fits, Cox’s Bazar (2021–2024).

**Table 3.**
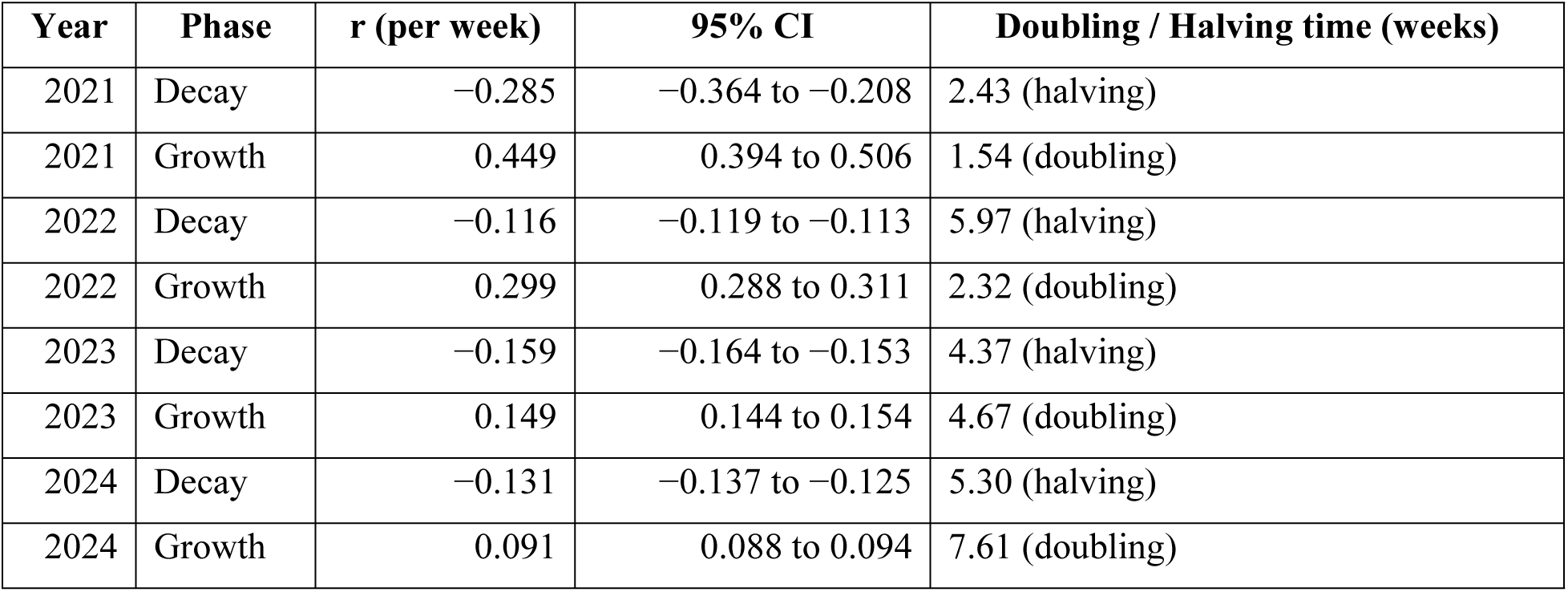
Phase-specific weekly epidemic growth and decay parameters from Poisson regression (2021–2024).

| Year | Phase | r (per week) | 95% CI | Doubling / Halving time (weeks) |
| --- | --- | --- | --- | --- |
| 2021 | Decay | −0.285 | −0.364 to −0.208 | 2.43 (halving) |
| 2021 | Growth | 0.449 | 0.394 to 0.506 | 1.54 (doubling) |
| 2022 | Decay | −0.116 | −0.119 to −0.113 | 5.97 (halving) |
| 2022 | Growth | 0.299 | 0.288 to 0.311 | 2.32 (doubling) |
| 2023 | Decay | −0.159 | −0.164 to −0.153 | 4.37 (halving) |
| 2023 | Growth | 0.149 | 0.144 to 0.154 | 4.67 (doubling) |
| 2024 | Decay | −0.131 | −0.137 to −0.125 | 5.30 (halving) |
| 2024 | Growth | 0.091 | 0.088 to 0.094 | 7.61 (doubling) |

### Clinical characteristics

Of the 35,581 cases reviewed, 8% (2,863 cases) necessitated inpatient admission. The majority, 90.6% (32,223 cases), were managed on an outpatient basis, while only 1.3% required referral to a tertiary facility. There were four reported deaths, resulting in a case fatality rate of less than 0.1%. (Table 4)

**Table 4.** Clinical profile and management outcomes among dengue cases across 2021-2024 (N = 35,581)

| A. Management and clinical outcome |  |
| --- | --- |
| Characteristic | n (%) |
| **Admitted** | 2,863 (8.0) |
| **Not admitted** | 32,718 (92.0) |
| OPD treatment | 32,223 (90.6) |
| Referred | 466 (1.3) |
| Discharged on same day | 21 (0.1) |
| Absconded | 4 (<0.1) |
| Died at home | 3 (<0.1) |
| Died in facility | 1 (<0.1) |

| **B. Disease severity (WHO classification)** |  |
| --- | --- |
| **Severity category** | **n (%)** |
| Group A: Mild dengue | 31,836 (89.5) |
| Group B: Dengue with warning signs | 3,587 (10.1) |
| Group C: Severe dengue | 158 (0.4) |
| **C. Clinical diagnosis** |  |
| **Clinical diagnosis** | **n (%)** |
| Asymptomatic dengue | 111 (0.3) |
| Dengue fever | 35,369 (99.4) |
| Dengue hemorrhagic fever | 35 (0.1) |
| Dengue shock syndrome | 60 (0.2) |
| Dengue with multiorgan failure | 6 (<0.1) |

Figure 4 illustrates the distribution of symptoms reported by the patients during their first presentation. Other than documented fever, dengue cases are most frequently presented with headache (58.7%) and malaise (55.1%). One third of cases presented with cough (30.8%). A substantial proportion of patients presented with gastrointestinal symptoms including nausea (18.8%), anorexia (18.4%), vomiting (14.0%) and abdominal pain (5.6%). Musculoskeletal symptoms included joint pain (16.3%) and body ache (3.8%). Retro-orbital pain was reported among 7.5% of the participants. Severe manifestations were very rarely reported, which include bleeding manifestation (0.3%), rash (0.1%), hypotension (0.1%) delirium (1.7%), respiratory distress (0.1%) and convulsion (<0.1%).

**Figure 4.**
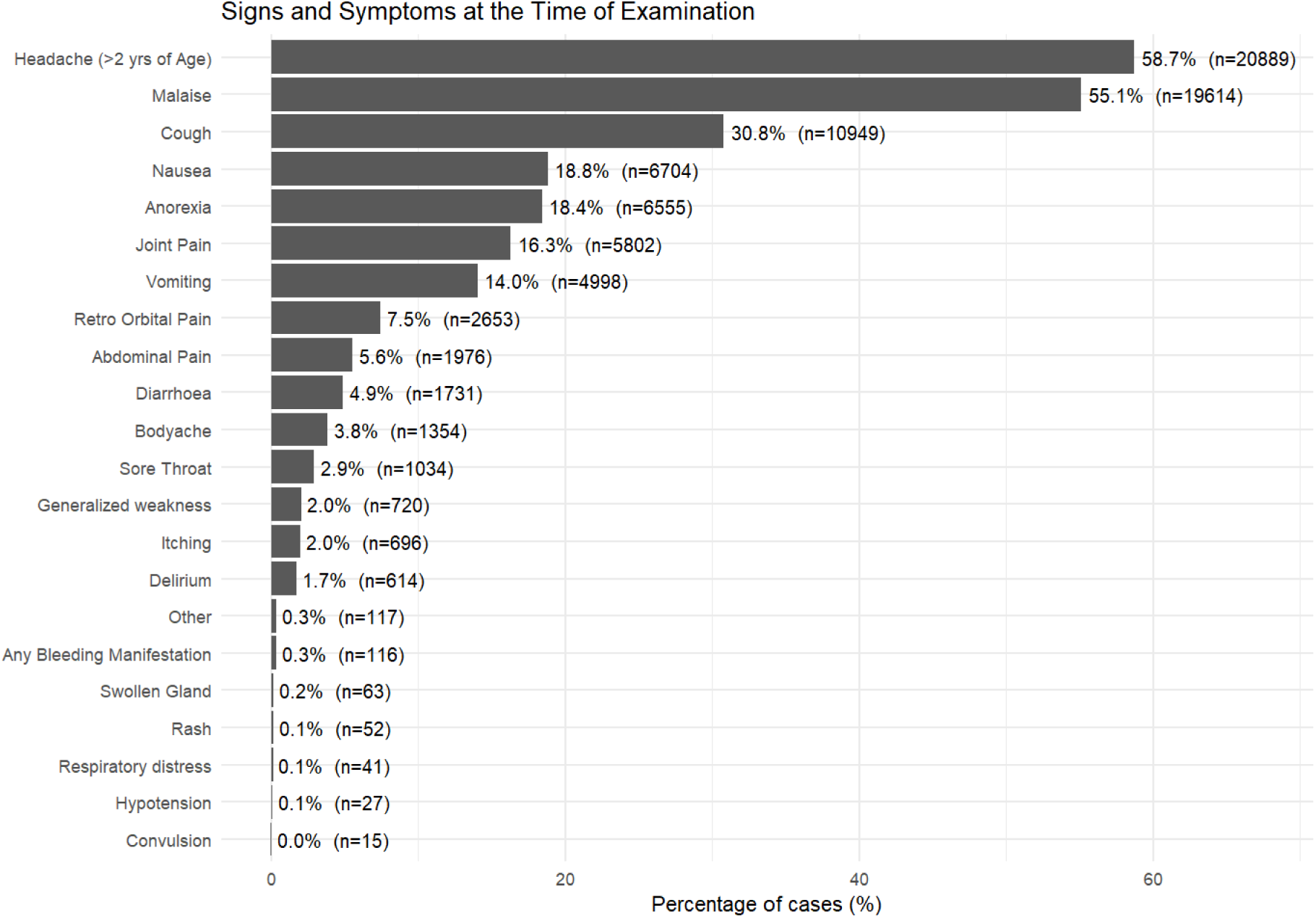
Frequency of symptoms reported at first clinical presentation among RDT-confirmed dengue cases (N = 35,581).

While over half of the patients had a recorded fever (≥38 °C), other vital signs of the majority of the patients stayed within normal ranges (Table 5). Low systolic BP (<90 mmHg) was observed in 2.8% of cases, and low diastolic BP (<60 mmHg) appeared in 6.4%. Furthermore, 4.6% of patients exhibited a prolonged capillary refill time (>2 seconds).

**Table 5.** Vital signs at presentation among dengue patients.

| Vital sign | Valid records (n) | Mean | Median | Mode | Observed range | Abnormal value (%) |
| --- | --- | --- | --- | --- | --- | --- |
| **Body temperature ($^{\circ}\text{C}$)** | 35,581 | 37.6 | 38 | 37 | 36–42 | Fever $\geq 38^{\circ}\text{C}$:<br>50.2% |
| **Systolic BP (mmHg)** | 18,864 | 108.3 | 109 | 100 | 40–196 | $<90$ mmHg:<br>2.8% |
| **Diastolic BP (mmHg)** | 18,864 | 69.1 | 70 | 70 | 30–140 | <60 mmHg: 6.4% |
| **Capillary refill time (sec)** | 35,581 | 1.55 | 2 | 1 | 1–4 | >2 sec: 4.6% |

WHO warning signs were observed in only a small proportion of patients as illustrated in Table 6. The most frequently reported warning signs included abdominal pain (3.6%), lethargy or restlessness (3.3%), and persistent vomiting (2.2%). Less commonly noted warning signs were present in fewer than 0.2% of cases, comprising of mucosal bleeding, clinical fluid accumulation, laboratory evidence of rising haematocrit, and liver enlargement exceeding 2 cm. Co-existing morbidities include infancy (2.9%) and old age (2.3%), pregnancy (1.5%) and chronic non-communicable diseases (Table 6).

**Table 6.** WHO warning signs and co-existing morbidities at presentation across 2021 to 2024 (N = 35,581)

| **WHO warning sign** | **n (%)** |
| --- | --- |
| **Abdominal pain** | 1,294 (3.6%) |
| **Lethargy / restlessness** | 1,181 (3.3%) |
| **Persistent vomiting** | 769 (2.2%) |
| **Mucosal bleeding** | 58 (0.2%) |
| **Clinical fluid accumulation** | 18 (0.1%) |
| **Laboratory: increase in haematocrit** | 13 (<0.1%) |
| **Liver enlargement >2 cm** | 11 (<0.1%) |
| **Co-existing morbidities** |  |
| **Extreme age** |  |
| **Infancy** | 1,031 (2.9%) |
| **Old age** | 801 (2.3%) |
| **Pregnancy** | 523 (1.5%) |
| **Chronic non-communicable diseases** |  |
| **Hypertension** | 200 (0.6%) |
| **Diabetes mellitus** | 162 (0.5%) |
| **Bronchial asthma** | 51 (0.1%) |
| **Heart disease** | 31 (0.1%) |
| **Stroke** | 13 (<0.1%) |
| **Chronic haemolytic anaemia** | 10 (<0.1%) |
| **Obesity** | 9 (<0.1%) |
| **Renal failure** | 4 (<0.1%) |

Multivariable logistic regression analysis demonstrates association of several demographic, clinical, and physiological factors with inpatient admission among patients with dengue in the refugee setting (Table 7). The likelihood of inpatient admission grew with age: individuals aged 15–29 had an adjusted odds ratio (aOR) of 1.61 (95% CI: 1.43–1.80), those aged 30–59 had an aOR of 1.93 (95% CI: 1.71–2.17), and people aged 60 or older had an aOR of 2.31 (95% CI: 1.90–2.81) when compared to children aged 0–14. Male patients have lesser likelihood to be admitted in comparison to females (aOR 0.88; 95% CI 0.80–0.96). Delay in presentation from the onset of symptoms have been found associated increase inpatient admission (aOR 1.06; 95% CI 1.04–1.08). Refugees had significantly higher odds of admission compared with host community patients (aOR 1.39; 95% CI 1.18–1.64). Clinically, the presence of any WHO dengue warning sign was strongly associated with hospital admission (aOR 26.60; 95% CI 24.15–29.30). Both low systolic BP (aOR 2.84; 95% CI 2.15–3.74) and low diastolic BP (aOR 1.60; 95% CI 1.29–1.98) were found associated with significantly higher odds of admission. Patients with any chronic co-morbidity had over sixfold increased risk of inpatient admission compared with those without comorbid conditions (aOR 6.07; 95% CI 4.71–7.82). The model demonstrates that fever ≥38°C at presentation, pregnancy or a previous history of dengue infection were not independently associated with hospital admission.

**Table 7.** Multivariable logistic regression predictors of inpatient admission among dengue patients cumulatively from 2021-2024.

| **Predictor** | **Adjusted OR (aOR)** | **95% CI** | ***p*-value** |
| --- | --- | --- | --- |
| Age group (years) |  |  |  |
| 15–29 vs 0–14 (ref) | 1.61 | 1.43–1.80 | <0.001 |
| 30–59 vs 0–14 (ref) | 1.93 | 1.71–2.17 | <0.001 |
| ≥60 vs 0–14 (ref) | 2.31 | 1.90–2.81 | <0.001 |
| Sex |  |  |  |
| Male vs Female (ref) | 0.88 | 0.80–0.96 | 0.005 |
| Delay in presentation (days) | 1.06 | 1.04–1.08 | <0.001 |
| Population type |  |  |  |
| Refugee vs Host (ref) | 1.39 | 1.18–1.64 | <0.001 |
| Fever ≥38 °C at presentation | 1.05 | 0.96–1.16 | 0.276 |
| Low systolic BP (<90 mmHg) | 2.84 | 2.15–3.74 | <0.001 |
| Low diastolic BP (<60 mmHg) | 1.6 | 1.29–1.98 | <0.001 |
| Any WHO warning sign present | 26.6 | 24.15–29.30 | <0.001 |
| Pregnancy | 1.01 | 0.70–1.45 | 0.973 |
| Any chronic co-morbidity | 6.07 | 4.71–7.82 | <0.001 |
| Previous dengue infection | 1.05 | 0.75–1.47 | 0.774 |

The model showed good discrimination (AUC = 0.82) and acceptable calibration across risk deciles, with no evidence of problematic multicollinearity.

### Exposure and behavioral characteristics

Table 8 summarizes the exposure history and behavioral characteristics of the dengue cases. Around 80% of the patients reported a history of increased mosquito bites in past three weeks and more than 60% reported the presence of stagnant water in their close surroundings. However, majority of the patients reported using at least one personal protective measure against mosquito bites. A small proportion (1.5%) of the patients reported a previous dengue episode.

**Table 8.** Exposure history and preventive factors among dengue cases (N = 35,581)

| Variable | Category | n (%) |
| --- | --- | --- |
| Previous dengue infection | No | 35,036 (98.5) |
|  | Yes | 545 (1.5) |
| Travel within 10 days before onset | No | 35,272 (99.1) |
|  | Yes | 309 (0.9) |
| Increased mosquito bites in past 3 weeks | No | 7,223 (20.3) |
|  | Yes | 28,358 (79.7) |
| Stagnant water in close surroundings | No | 13,627 (38.3) |
|  | Yes | 21,954 (61.7) |
| Family member with similar symptoms (past 10 days) | No | 32,155 (90.4) |
|  | Yes | 3,426 (9.6) |
| Use of protective measures | No | 4,273 (12.0) |
|  | Yes | 31,308 (88.0) |

### Diagnostic characteristics

Among the total pool of dengue patients, NS1 antigen alone was positive in 62.1%, IgM alone was positive in 33.6%, and both NS1 antigen and IgM/IgG were positive in 4.3% of the cases. Table 9 indicates a correlation between the delay from symptom onset to clinical presentation and RDT positivity patterns. Patients exhibiting NS1 antigen positivity exclusively reported the shortest interval before seeking medical care (mean 2.4 days, median 2 days). Those positive for both NS1 antigen and IgM/IgG antibodies experienced an intermediate delay (mean 4.1 days, median 4 days), while the longest delay was noted among patients positive solely for IgM/IgG antibodies (mean 5.0 days, median 5 days). After adjusting for age and sex, each additional day of delay from symptom onset to presentation was independently associated with higher odds of being IgM-only positive (aOR 2.43, 95% CI 2.38–2.47; p<0.001).

**Table 9.** Delay in presentation by RDT positivity pattern (N = 35,581)

| RDT positivity pattern | n (%) | Mean delay (days) | Median delay (days) | Kruskal–Wallis test |
| --- | --- | --- | --- | --- |
| NS1 antigen only positive | 22,082 (62.1) | 2.4 | 2 | $\chi^2 = 12,319$,<br>df = 2, $p < 0.001$ |
| Both NS1 antigen and IgM/IgG positive | 1,544 (4.3) | 4.1 | 4 |  |
| IgM/IgG only positive | 11,955 (33.6) | 5 | 5 |  |
| **Total** | **35,581 (100)** | — | — |  |

## DISCUSSION

According to the authors’ knowledge, this is the largest outbreak of dengue recorded in the refugee or displacement setting globally (1). This retrospective analysis confirms the exceptionally high burden of dengue among the Rohingya refugees in Cox’s Bazar with a caseload of more than 35,000 from 2021 to 2024. Extremely crowded living conditions with fragile and crowded shelters may contribute to the high transmission in the refugee setting as this provides abundance of human hosts for mosquitoes (11). Poor sewerage systems, stagnation of water near the households and poor waste management system may further contribute to the transmission by increasing the habitats for mosquitoes (7,26). This is further evidenced by finding of this study, as more than 60% of participants reported the presence of stagnant water in their close surroundings (Table 8). These findings indicate that dengue prevention strategies in the Rohingya camps must prioritize environmental vector control and improved water, sanitation, and waste management systems through stronger coordination between health, WASH, and camp management sectors.

Our study identified a significant seasonal increase in dengue cases, with peaks observed between weeks 25 and 38, corresponding to the late-and post-monsoon period. This pattern suggests that water stagnation following rainfall contributes to mosquito breeding and subsequent dengue transmission (11). The analysis indicates that dengue emerged as an acute outbreak in 2021, followed by larger-scale outbreaks with progressively slower growth rates in subsequent years. This finding may be due to the gradual shift of dengue hotspots to additional camps beyond the original sites where preventive measures were implemented. Apart from the seasonal peaks, low-level transmission throughout the year also confirms the disease is taking endemic form with partial population immunity and ongoing vector presence as reported in WHO’s entomological studies (11). We have also observed consistent shifting of peak periods across the years with a consistent halving time. While this points to the possible linkage to climate factors such as rainfall, humidity, and temperature, it also alarms the potentiality of sustained year-round outbreaks, which has already been documented in the broader context of Bangladesh(18,27). Therefore, there is critical need for shifting priorities from short-term outbreak response towards the sustained dengue preparedness, surveillance, and vector control. From a policy perspective, this transition toward sustained transmission requires integrating dengue preparedness into routine health system planning, including year-round surveillance, vector monitoring, and contingency resource allocation within humanitarian health programmes. This epidemiological transition suggests that dengue control strategies in the Rohingya camps should shift from short-term outbreak response toward sustained preparedness measures, including year-round surveillance, proactive vector control, and long-term programme planning.

We have found that the males accounted for a higher proportion of cases than females. This is consistent with the findings in other settings like the countries in South and South-east Asia, including Dhaka, Bangladesh (16). While the researchers yet could not conclude the underlying cause of the male predominance of dengue, in our setting, it could be related with the fact that men culturally spend more time engaged in outside activities and occupations that increase their likelihood to get exposure (17).

The study reported the spectrum of presenting symptoms for patients with dengue. While this study reported a significant proportion of common dengue symptoms, atypical symptoms such as cough, abdominal pain and diarrhea were also observed across all four years (12,21). Previous studies in urban settings in Bangladesh suggest that Bangladeshi healthcare workers have documented an increasing trend in gastrointestinal symptoms and a decreasing trend in classical hemorrhagic manifestations (like skin petechiae or melena) and joint pain in dengue cases (16), emphasizing the need for clinicians to maintain a high index of suspicion for dengue even in patients presenting with non-classical symptoms, particularly in high-transmission and resource-limited settings such as refugee camps. For clinical practice and programme planning, this highlights the need to strengthen clinician awareness across healthcare facilities and ensure that up-to-date diagnostic triaging protocols for dengue are available.

Overall, the study reported an extremely low fatality rate (case fatality 0.011% (<0.1%) and minimal severity among dengue cases: 10% of patients exhibited warning signs, only 0.4% were classified as severe dengue, and 8% required hospitalization. This could be attributed to the dengue response model implemented by IOM at primary healthcare setting incorporating early diagnosis by making diagnostic kits available at primary care facilities, linking the community healthcare workers in follow-up and training the clinicians for proper management. Early detection, timely surveillance, and prompt clinical consultation play a critical role in reducing dengue severity and mortality by enabling early case management, rapid identification of outbreaks, and timely referral of high-risk patients (11). These findings suggest that maintaining decentralized diagnostic capacity, standardized clinical protocols, and community-based follow-up systems should remain central components of dengue control policies in humanitarian health settings.

However, this assessment found only 1.5% of the reported cases to have a history of previous dengue infections, representing majority of these cases as a primary infection. Evidence suggests that subsequent dengue infections increase the risk of severe disease compared with primary infections due to mechanisms such as antibody-dependent enhancement (ADE), particularly when reinfection is caused by a different dengue virus serotype (10). Considering high population density, favorable climate for vector proliferation, inadequate vector control measures in the Rohingya refugee camps, there is a heightened likelihood of repeated exposure and secondary infections in the future, which may consequently increase the risk of severe dengue cases if preventive and surveillance measures are not strengthened (17). Policy planning should therefore prioritize sustained vector control, strengthened surveillance systems, and preparedness for potential increases in severe dengue associated with secondary infections.

Multivariable regression analysis of the study revealed that in this refugee setting, inpatient admission for dengue was primarily associated with age, delayed presentation, presence of warning signs, hypotension and presence of chronic co-morbidity rather than by fever alone or prior dengue exposure. Delayed presentation exposes patients to the critical phase of dengue, during which plasma leakage, hypotension, and organ dysfunction are more likely to occur, thereby increasing the need for inpatient admission; this relationship has been consistently reported in clinical and epidemiological studies from dengue-endemic, resource-limited settings(11,28,29). Therefore, risk communication and community engagement should be strengthened for increasing health seeking behavior, community-based referrals and rapid triage to prevent delays in presentation. This is consistent with earlier studies from the Rohingya camps that documented challenges in community engagement, preventive practices, and trust-building during infectious disease responses, underscoring the importance of strengthening community-based communication strategies in dengue control efforts (5,6).The strong association between hospital admission and warning signs confirms programme’s strong adherence to as well as high practicality of WHO warning signs and severity criteria for hospital admission prioritizing the high-risk groups. Strengthening community-based early detection and referral systems may therefore reduce delays in care-seeking and prevent progression to severe disease requiring hospitalization.

The study confirmed predominant NS1 antigen positivity among patients presenting early after symptom onset and IgM antibody positivity among those presenting later, with a brief overlapping period during which both markers were detectable. This pattern aligns with the pathophysiology of dengue infection, in which NS1 antigenemia is highest during the early febrile phase (typically within the first 3–5 days of illness) and declines as viremia resolves, while dengue-specific IgM antibodies become detectable from approximately day 4–5 onward and persist into the convalescent phase. Consequently, in humanitarian settings where delayed care-seeking is common, reliance on NS1-based testing alone would fail to identify a substantial proportion of dengue cases. The use of IgM/IgG rapid diagnostic tests during the convalescent phase is therefore essential for confirming dengue infection. This finding is consistent with current World Health Organization and Centers for Disease Control and Prevention guidance, which recommend serological testing for patients presenting beyond the early febrile phase of illness (11,30–32). Ensuring the availability of both NS1 antigen and IgM/IgG rapid diagnostic tests across primary healthcare facilities is therefore essential for accurate case detection and appropriate clinical management in humanitarian settings.

The findings from this research provide several important policy insights. The shift from episodic to sustained dengue transmission in Cox’s Bazar indicates the need for shift from emergency response to long-term preparedness. The delayed care-seeking and diagnosis indicates the importance of strengthening surveillance, risk communication and community engagement, referral, and diagnostic services at primary health facilities. Furthermore, the findings from the environmental and exposure factors indicate the need for better coordination between sectors to provide better vector and environmental management. Overall, these findings indicate that strengthening these measures are essential to reducing dengue transmission, promoting early presentation to health facilities and ultimately, improve the morbidity and mortality outcomes among dengue patients.

## CONCLUSION

The study documented the epidemiological, clinical and diagnostic characteristics of the largest documented dengue outbreak in a refugee or forced displacement setting globally. Our findings indicate that dengue transmission in the Rohingya refugee camps is no longer a short-lived outbreak phenomenon but is evolving toward sustained transmission with the potential for increasing severity in future outbreaks. The combination of high population density, environmental suitability for vector breeding, delayed care-seeking, and a largely immunologically naïve population underscores the urgent need to shift from episodic outbreak response to a long-term dengue control strategy. This should include sustained vector control and environmental management, strengthened routine and syndromic surveillance, integrated use of NS1 and IgM/IgG diagnostics across all phases of illness, and continued investment in community-based risk communication, early referral, and triage systems. Proactive identification and close monitoring of high-risk groups — particularly older adults, patients with co-morbidities, and those presenting late — will be critical to preventing progression to severe disease. Strengthening these components within humanitarian health systems is essential to mitigating the future burden of dengue in protracted displacement settings such as Cox’s Bazar.

## DATA AVAILABILITY

The data underlying the findings of this study are available from Zenodo at DOI: https://doi.org/10.5281/zenodo.19219551. The dataset includes anonymized and aggregated data used in the analysis.

## ACKNOWLEDGEMENTS

CEH led the study conceptualization and design, developed the protocol, oversaw the overall methodology, and led the analysis and drafting of the manuscript. PPD and MMR contributed to data cleaning and epidemiological/statistical analysis, and supported interpretation of findings. MAH led data verification and quality checks, supported data cleaning, and conducted manuscript coherence review and proofreading. SH and SHS supported data compilation and cleaning. MAT supported field coordination and implementation follow-up. MAH contributed to analysis and supported manuscript review. ERS and JCO provided research supervision and programme-level oversight to ensure alignment with operational priorities. UMP and DB contributed to technical review and refinement of the manuscript. JPA provided technical input through the Global Migration Health Research & Epidemiology Unit. DP and SMNM contributed to analysis support and technical review. MH and ATMHB supported coordination, review, and research uptake with relevant authorities, and MFH provided technical review and supported research uptake. All authors reviewed and approved the final version of the manuscript. CEH is the corresponding author and JCO is the guarantor of the study.

## GENERATIVE AI DISCLOSURE

Language editing assistance was provided using Grammarly and ChatGPT (OpenAI; GPT-5.2 Thinking, accessed 08 February 2026), limited strictly to grammar, clarity, and sentence-level flow. No new scientific content, analyses, results, figures, or references were generated. All authors reviewed and approved the final manuscript and take full responsibility for the accuracy and integrity of the work.

## SUPPORTING INFORMATION

**S1 Figure. RDT positivity patterns by delay from symptom onset to presentation.** Distribution of NS1 antigen–only, IgM/IgG–only, and dual RDT positivity patterns according to delay in days from symptom onset to presentation among dengue cases.

**S2 File. Dengue case report form used for routine outbreak surveillance and clinical data collection.** Standardized case reporting form capturing demographic, epidemiological, clinical, laboratory, and outcome variables for dengue patients across IOM-supported health facilities.

